# Seasonal influenza vaccination in people who have contact with birds

**DOI:** 10.1101/2024.11.03.24316665

**Authors:** Amy Thomas, Suzanne Gokool, Harry Whitlow, Genevieve Clapp, Peter Moore, Richard Puleston, Louise E Smith, Riinu Pae, Ellen Brooks-Pollock

## Abstract

**Background:** Following the 2021-2022 avian influenza panzootic in birds and wildlife, seasonal influenza vaccines have been advised to occupationally high-risk groups to reduce the likelihood of coincidental infection in humans with both seasonal and avian influenza A viruses.

**Methods:** We developed and launched a questionnaire aimed at poultry workers and people in direct contact with birds to understand awareness and uptake of seasonal influenza vaccination. We collected responses in-person at an agricultural trade event and online.

**Findings:** The questionnaire was completed by 225 individuals from across the UK. The most commonly reported reason for vaccination was protection against seasonal influenza (82%, 63 of 77). Nearly all individuals aged ≥65 years reported that the vaccine was recommended for them (24 of 28). There was no difference in recommendation for occupational groups. Most vaccinees were aged over 60 years (60%, 29 of 48), however coverage was lower than expected in the ≥65 target group. Vaccination in those exposed to avian influenza was low (32%, 9 of 28). Not having enough time was the single most reported reason for not getting vaccinated in those intending to. Individuals unintending to be vaccinated perceived natural immunity to be better than receiving the vaccine as well as lack of awareness and time.

**Conclusions:** Our findings suggest that targeted campaigns in occupationally exposed groups need to be undertaken to improve communication of information and access to vaccine clinics. We recommend co-production methods to optimise this public health strategy for increased knowledge and future vaccine uptake.

## Introduction

Influenza is a globally important pathogen with a high pandemic potential. Of the four influenza virus types, influenza A (subtypes H1N1, H2N2 and H3N2) and B circulate in the human population and cause seasonal influenza epidemics, usually in the winter in temperate regions^1^. The high pandemic potential of influenza is due to the ability of distinct subtypes to reassort either in human or porcine hosts and create novel influenza subtypes, against which there is little or non-existing prior immunity in the human population (antigenic shift)^2, 3^. Recent increases in avian influenza (AI) outbreaks in Europe and America have heightened concern about the risk of a novel virus emerging.

Wild birds, primarily waterfowl, are considered a natural reservoir for influenza A viruses, where all subtypes circulate in the low pathogenic form, causing limited or no symptoms^4^. However, influenza A subtypes (H5 and H7) are considered Highly Pathogenic AI (HPAI) because of rapid transmission, serious infection and death in domestic and wild birds^5,6^. In recent years, there has been an increase in HPAI A(H5N1) influenza outbreaks in wild and domestic birds. In 2021-2022, 48 million birds were culled due to HPAI across Europe, with many further deaths in wild birds from infection^7^. Transmission of HPAI A(H5N1) clade 2.3.4.4b in wild birds continued throughout summer 2022 in Great Britain and northwest Europe, unlike previous seasons^8^. Furthermore, there have been increased reports of infection in wild mammals during this panzootic, with evidence of genomic changes to support an advantage for mammalian infection and transmission^9^. In the USA, recent widespread transmission in cattle and isolated cases of cattle-to-human transmission has raised concerns that further transmission could occur to humans, leading to epidemics or a pandemic and resulting in heightened public health risk assessments^10,11,12^.

Most cases of HPAI in humans occur following very close contact with infected birds and ranges in severity. Recent cases have mostly been asymptomatic or mild and self-limiting. However, infection with other strains has been serious or fatal^13^. In the UK, public health action is initiated for suspected or confirmed AI infections in birds to prevent human infection, including advice and training on appropriate use of PPE, information about HPAI, and possible follow up and prophylactic antiviral treatment for people who have been exposed^14^. An asymptomatic screening programme of 144 exposed persons identified four H5N1 human infections^13,14^.

In the UK, seasonal influenza vaccination is recommended for adults over 65 years, pregnant women, individuals with certain long-term health conditions and those caring for people with a weakened immune system, and this is similar to many European countries^17^. Some European countries recommend seasonal influenza vaccination for occupationally exposed groups and to other groups exposed to animals potentially infected with avian influenza, with the aim of minimising human influenza and avian influenza co-infection risk, subsequently reducing reassortment events^18^. In 2023 in the UK, the Joint Committee on Vaccination and Immunisation (JCVI) advised that workers in the poultry and avian animal health industries should be considered for the seasonal inactivated influenza vaccine^19^. Individuals highlighted at greatest risk included those undertaking culling or cleaning at confirmed avian influenza outbreak premises, handling live unwell birds, those with direct exposure to bird faeces/litter through initial egg sorting or cleaning and those collecting suspected/confirmed infected wild bird carcasses^19^. Notably, seasonal influenza vaccines do not provide specific protection against avian influenza but there may be some level of cross-protective immunity via cross-reactive neutralising antibody and T-cell responses^20^.

Here, we report on awareness and attitudes to seasonal influenza vaccination amongst people who have contact with birds in the UK. We used data from the Avian Contact Study, a co-developed questionnaire aimed at poultry workers and individuals in contact with any type of bird(s). Our findings aim to inform discourse on including this high-risk group in future seasonal influenza vaccination programmes.

## Methods

### Brief description of the Avian Contact Study

The Avian Contact Study aimed to inform and optimise current zoonotic influenza public health measures. A survey was co-produced with members of the poultry industry, UK Health Security Agency, Animal Plant Health Agency and the Health Protection Research Unit (HPRU) in Behavioural Science and Evaluation to understand awareness and potential transmission of HPAI among people who have contact with domestic or wild birds such as poultry farmers and bird-keepers. A questionnaire was developed and deployed to collect information on the type of contact people have with birds, their social contacts, perceptions of avian influenza generally and perceptions of their own personal health. Full details of the questionnaire methodology can be found in the Data Note^21^ and a pdf of the questionnaire is available as *Extended data*^22^.

### Ethics and consent

Ethical approval for the study was obtained from the University of Bristol, Faculty Research Ethics Committee, approval number 17048 on 16 January 2024.

Informed written consent (using e-consent hosted on Research Electronic Data CAPture tools, REDCap) for the use of data collected via the questionnaire was obtained from respondents.

### Data collection

The questionnaire was initially delivered in person at the British Pig and Poultry Fair (15th-16th May 2024) and was subsequently circulated online via various poultry forums and networks via word-of-mouth. All questions were optional. Data captured between 15 May 2024 and 31 July 2024 were included in these analyses. Questionnaire data were collected using REDCap^23^, hosted at the University of Bristol. Data were exported from REDCap to R studio (version 4.4.0) for data manipulation and analysis.

### Demographic and background questions

We collected background demographic data on age, gender, occupation and current levels of health.

### Influenza vaccination

The following vaccine-related questions were included in the Avian Contact Study questionnaire:

1. Has the seasonal (winter) flu vaccine provided by the NHS been recommended for you?
  º Yes/No/I don’t know
2. Have you been vaccinated against seasonal (winter) flu in the last 12 months?
  º Yes as provided by the NHS
  º Yes, I paid for it myself;
  º No, but I intend to get vaccinated;
  º No, I am undecided;
  º No, I am not intending to get vaccinated;
  º I do not know.
3. [If vaccinated] When were you vaccinated?
4. [If vaccinated] What factors influenced your decision to get vaccinated?
  º For protection against seasonal (winter) flu
  º Convenience of appointment times
  º Convenience of vaccine centre location
  º The vaccine is safe – no side effects
  º The vaccine is effective at reducing the risk of seasonal (winter) flu
  º Other people I know are vaccinated
  º The vaccine has been recommended for me
  º For protection against avian influenza (bird flu)
  º To protect other people
5. [If not vaccinated/not planning to get vaccinated/undecided] What factors have prevented you from getting vaccinated?
  º The vaccine is too expensive
  º I am not sure if the vaccine is effective
  º The vaccine may cause short term side effects
  º The vaccine may cause long term side effects
  º I am not likely to catch seasonal (winter) flu
  º I will not get very ill if I catch seasonal (winter) flu
  º I am allergic to the vaccine
  º It is better to get natural immunity
  º Other

### Data analysis

Records with responses for at least one of the five vaccine questions were retained for analysis. The proportion of individuals responding to each question was calculated and reported alongside 95% confidence intervals for a single proportion using the two proportions z-test, as well as raw counts. Responses were grouped by age, gender and occupation. Occupations with fewer than 5 individuals per category are not shown to preserve anonymity. We calculated seasonal influenza vaccine uptake for individuals based on their eligibility to receive the vaccine via the NHS guidance of individuals ≥65 years old. We further considered an ‘at-risk’ group defined as individuals reporting exposure to AI virus. We could not reliably identify occupationally at-risk individuals as per the Green Book guidance (poultry and related workers) due to ambiguity in occupational risk^19^. Individuals <65 years old and who did not report AI virus exposure were grouped as ‘no risk’. We grouped responses for question 2 (vaccination status) into four categories: Yes (Yes as provided by the NHS or Yes I paid for it myself); No (No I am not intending to get vaccinated); Intending (No but I intend to get vaccinated); and Don’t know (I do not know or No I am undecided).

## Results

A complete description of methods, public involvement and participant demographics can be found in the Data Note^21^. A total of 225 people took part in the study between May to July 2024, two respondents were excluded from all analyses as they did not provide responses to at least one vaccine question. Briefly, most respondents were male (63%, 140/222) and over 30 years (81/222, 36%) (Note one participant did not complete the gender question). Male respondents had a higher median age than female respondents (52 compared to 44 years) (Figure 1a). Poultry farmer was the most frequently reported occupation (102/222, 72%), followed by veterinarian, zookeeper and poultry manager (Figure 1b).

**Figure 1:**
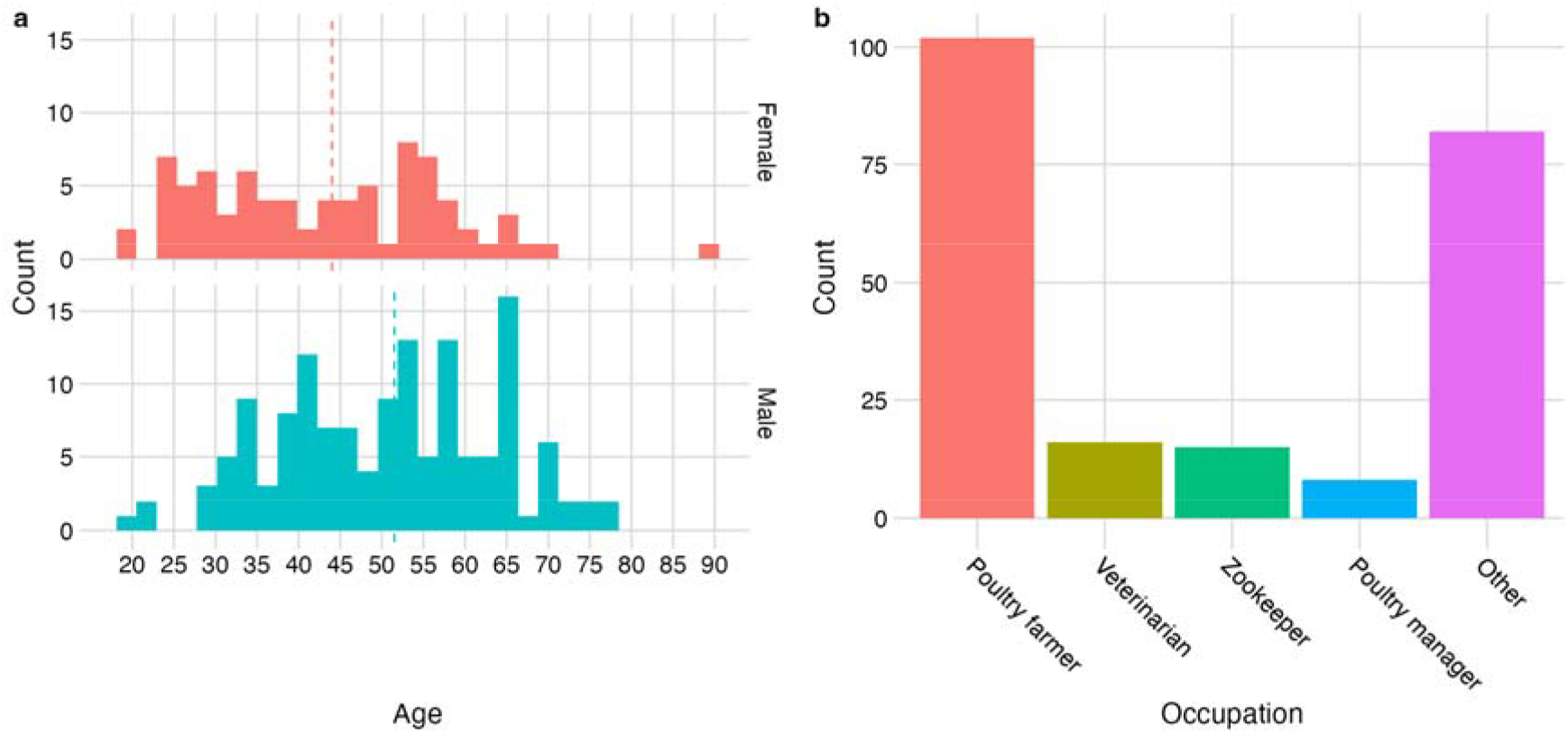
Distribution of respondent age, gender and occupation. **a**, Age distribution of respondents grouped by gender – female in red and male in blue (note one ‘Other’ and one NA response not plotted. **b**, Top 4 occupations reported (occupations with fewer than 5 observations grouped with ‘Other’).

### Seasonal influenza vaccination

#### Eligibility for free seasonal influenza vaccination

Across all age groups, 52% (116 of 222) of individuals reported that the seasonal influenza vaccine was recommended for them, 4% did not know. The proportion of individuals reporting recommendation increased with age (Figure 2a). Nearly all individuals aged 65 years and over reported that the vaccine had been recommended for them (86%, 24 of 28) but less than half of under 65’s reported recommendation (47%, 92 of 195). There was no evidence of a difference in recommendation among commonly worked occupations (Figure 2b). Taken together, this suggests low awareness of the vaccine eligibility criteria for workers in the poultry and avian animal health industries to receive seasonal influenza vaccine.

**Figure 2:**
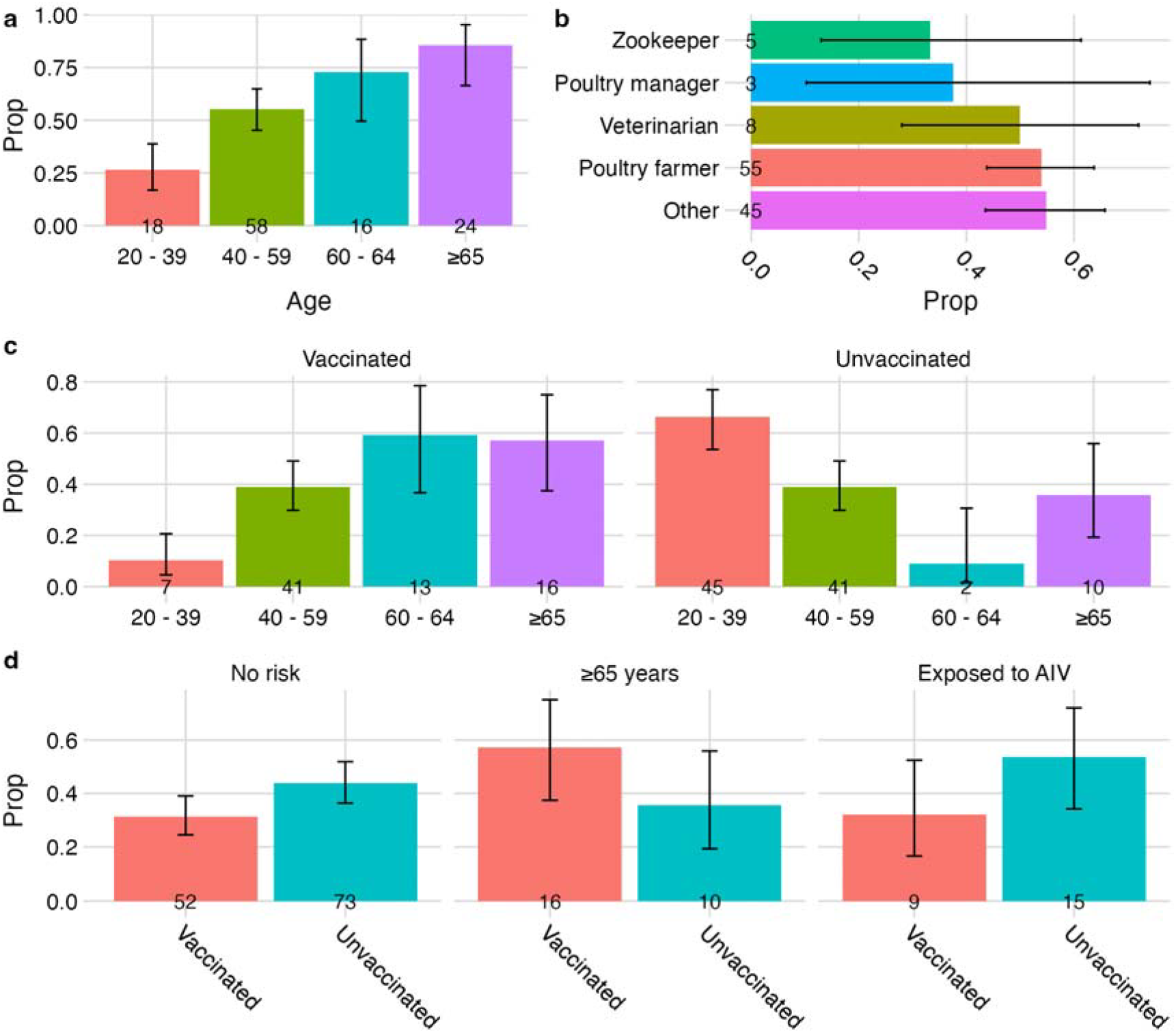
Seasonal influenza vaccine recommendation and uptake. **a**, Proportion self-reporting recommendation to receive seasonal influenza vaccine by age. **b**, Proportion self-reporting recommendation to receive seasonal influenza vaccine by occupation. **c**, Proportion of individuals receiving seasonal influenza vaccine in the last 12 months by age. **d**, Proportion of individuals receiving seasonal influenza vaccine in the last 12 months by eligibility/risk group. Those considered ‘at-risk’ reported being exposed to avian influenza virus (AIV). Error bars represent 95% confidence intervals for single proportion.

#### Trends in seasonal influenza vaccine uptake

Overall, 35% of respondents (77 of 223) had been vaccinated against seasonal influenza, of these 79% (61 of 77) had the vaccine provided by the NHS and a further 21% paid for the vaccine themselves (16 of 77). Most individuals did not intend to receive the vaccine (44%, 98 of 223) whilst 11% (25 of 223) had not been vaccinated but intended to and 10% (22 of 223) did not know. Seasonal influenza vaccine uptake increased with increasing age; the greatest proportion of vaccinees (60%, 29 of 48) were aged ≥60 years (Figure 2c). Conversely, younger age groups were less likely to be vaccinated, with the largest unvaccinated proportion (66%, 45 of 68) aged between 20-39 years (Figure 2c).

We considered vaccine uptake based on eligibility and risk. Given our sample, 28 of 223 respondents were ≥65 years old and eligible to receive seasonal influenza vaccine as per the UK seasonal influenza vaccine campaign17.Of these, just over half had been vaccinated in the previous 12 months (57%, 16 of 28). Vaccine uptake in those reporting avian influenza virus exposure (32%, 9 of 28) was lower than the ≥65-year group but marginally higher than the no-risk group (31%, 52 of 166) (Figure 2d).

#### Reasons for seasonal influenza vaccination

Among those who were vaccinated, the most commonly reported reason for vaccination was protection against seasonal influenza (82%, 63 of 77). Protection against avian influenza, convenience of getting vaccinated (location/appointment time) and the behaviour of others were infrequently reported as reasons for being vaccinated (Figure 3a).

**Figure 3:**
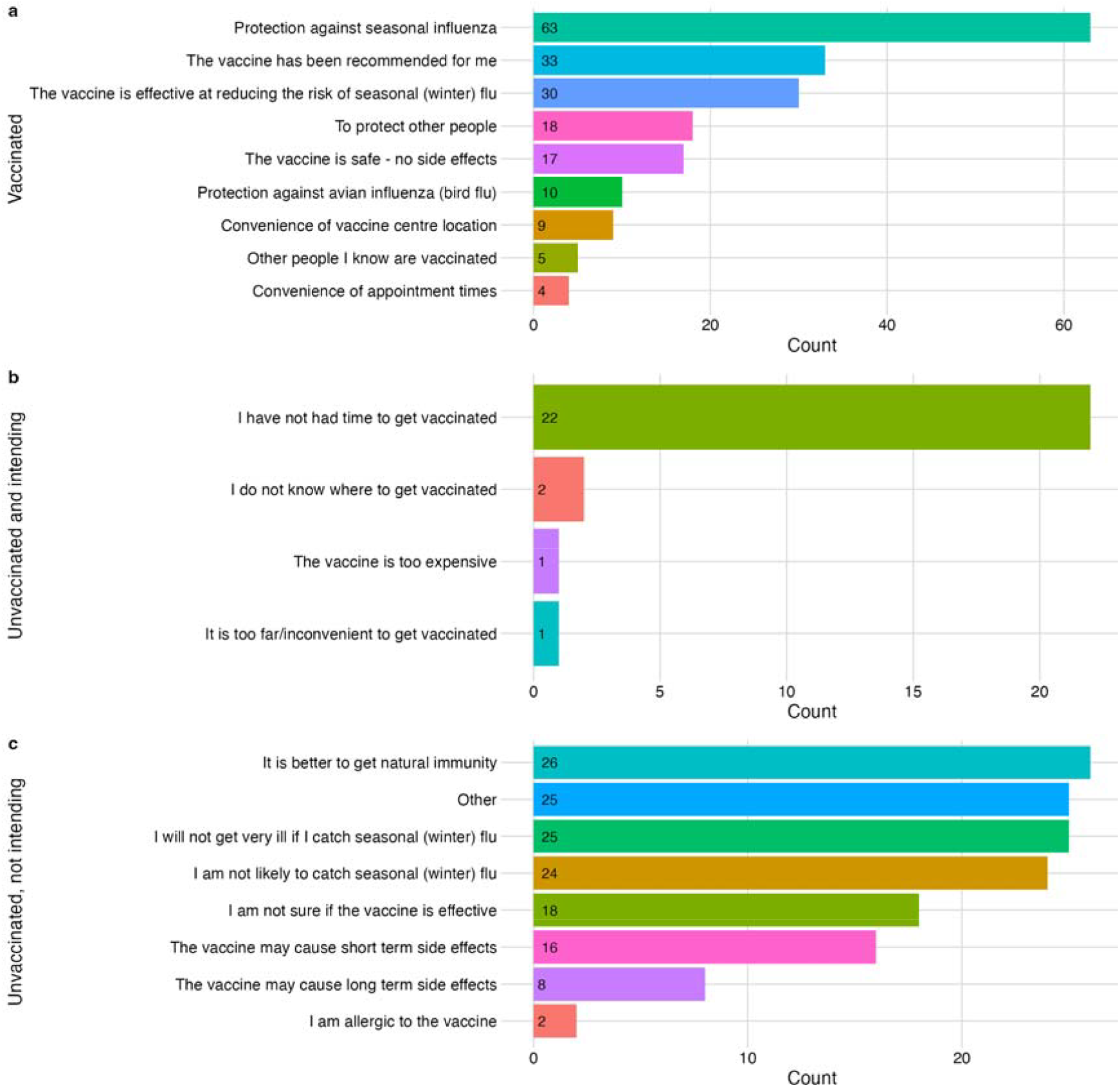
Reasons for seasonal influenza vaccination. **a**, reasons for receiving the seasonal influenza vaccine among vaccinated individuals, **b**, reasons preventing an individual from becoming vaccinated among individuals intending to be vaccinated, **c**, factors influencing indecision or decision to not be vaccinated among those not intending to be vaccinated.

Amongst individuals who intended to get vaccinated, not having enough time to get vaccinated was the single most reported reason for not having the seasonal influenza vaccine (88%, 22 of 25; Figure 3b).

Amongst individuals who were not intending to get vaccinated, frequently reported reasons for not getting vaccinated were perceiving that it was better to get natural immunity than to receive the vaccine (27%, 26 of 98), and thinking that the seasonal influenza would not make them very ill if they did catch it. Short or long-term side effects were less frequently reported reasons. Of the responses given as free-text for ‘Other’, lack of awareness and lack of time were commonly reported reasons for not becoming vaccinated (Figure 3c).

## Discussion

Seasonal influenza vaccination is an important public health measure aimed at limiting influenza-related morbidity and mortality. Although not intended for protection against avian influenza infection, recent increases in avian influenza in birds and mammals have increased the risk of human-avian influenza co-infection and reassortment. As a result, poultry keepers and others at high risk of avian influenza infection have become eligible in seasonal influenza vaccine programmes. The analysis presented here suggests that awareness and uptake in individuals at high risk of avian influenza exposure is low.

Self-reported reasons for not being vaccinated, including not having time or knowing where to get vaccinated, highlighting the need for accessible and convenient vaccine centres, especially in rural areas. The Fit2Farm survey found that UK farmers work long hours and have limited days off ^24^, therefore accessing vaccine centres may be challenging. Efforts to improve access to healthcare for farmers include outreach programmes and nurse-led services. One example is a pop-up site where the influenza vaccine was available^25^. Our results support the need for flexible vaccine delivery models that are tailored to meet the demands of individuals working in poultry and farming related25,26. The importance of these agile systems has also been highlighted for delivery of antivirals to those exposed to avian influenza^28^.

The WHO and EU target for influenza vaccine coverage rate is 75% in target groups. In our sample, those receiving the vaccine were mostly older individuals aged ≥60 years from the NHS to prevent them from seasonal influenza (60%). Since 2020, the estimated uptake in individuals aged 65 and over in England is between 78% and 82%. Prior to the COVID pandemic, uptake ranged between 70% and 74%^29^. Here, we report lower coverage, with just over half of target individuals aged ≥65 years (57%) having been vaccinated, albeit the sample size is small, reflecting the need to create vaccine strategies that are suitable to this demographic.

We identified a miscommunication or lack of understanding regarding why individuals in contact with birds are recommended to receive seasonal influenza vaccine. Non-vaccinated individuals reported that they thought it was better to obtain natural immunity, and protection against avian influenza was not a commonly cited reason for vaccination (the rationale for vaccine consideration for this group was to prevent reassortment of the virus). Furthermore, participants who come into occupational contact with poultry were not aware of the advice for seasonal influenza vaccination from the 2023 Joint Committee on Vaccination and Immunisation (JCVI). This highlights the need to enhance engagement activities with this group for improved communication in developing and delivering vaccination campaigns. Co-production and co-creating methodologies can be effective in engaging with members of the farming community to undertake zoonotic disease research^30^ and can offer a method for co-producing interventions with the community^31^.

This questionnaire provides a timely and current snapshot of attitudes and uptake relevant for planning seasonal autumn influenza vaccine campaigns for people in contact with poultry. Moreover, our approach provides information for members of the farming community who have historically been identified as vulnerable in terms of health and occupational needs^32^ as well as experiencing barriers with primary health care providers^33^. We achieved good coverage of respondents in older age categories through in-person and online questionnaire completion^21^. Our sample was composed of mostly poultry farmers and hobby bird-keepers and those in contact with wild birds are underrepresented, therefore generalisation to other groups in contact with birds should be done cautiously. In-person recruitment at an agricultural show may have excluded ‘front-line’ poultry workers due to preferential attendance from managers and specialists in this field. Further, the questionnaire and promotion were conducted in English, possibly marginalising groups – many poultry subcontractors do not have English as their first language^34^. Reporting biases may also exist between online and in-person responses. Interpretation of small sample sizes should be done with caution. Future studies should seek to address these data gaps by maximising recruitment across multiple groups and sectors.

## Conclusion

We have highlighted the disconnect between policy and influenza vaccine implementation. There is a need to work with members of the poultry and wild-bird community for informing public health responses. Current recommendations to vaccinate this cohort will be thwarted if tailored campaigns are not undertaken. We recommend using co-production to identify methods for improving access to vaccine clinics and explore ways for effective communication.

## Data Availability

Repository data.bris: The Avian Contact Study: questionnaire data 15 May to 31 July 2024 Due to the sensitivity of the data involved, these data are published as a restricted dataset at the University of Bristol Research Data Repository data.bris, at https://doi.org/10.5523/bris.3nmqsrbv5ruom2abn0ql6e8yh235. The metadata record published openly by the repository at this location clearly states how data can be accessed by bona fide researchers. Requests for access will be considered by the University of Bristol Research Data Service, who will assess the motives of potential data re users before deciding to grant access to the data. No authentic request for access will be refused and re users will not be charged for any part of this process.
This project contains the following underlying data:
Data file 1. (Raw underlying questionnaire data, csv file)
Data file 2. (Raw underlying questionnaire data, .RDS file)
Data file 2. (Associated data dictionary, csv file)
Data file 3. (Code for importing underlying data in csv format into R for setting up labelled data, .r file)
Data file 4. (Blank consent form and participant information sheet, pdf file)
Data are available under the terms of National Archives Non Commercial Government Licence for public sector information.
Extended data
Repository Zenodo: The Avian Contact Study Questionnaire and Data Dictionary [10.5281/zenodo.13617061]
This project contains the following extended data:
AvianInfluenzaSocialContactSu.pdf (The final questionnaire REDCap, PDF)
AvianInfluenzaSocialContactSur_DataDictionary_080824v1.csv (Associated data dictionary, csv file)
Data are available under the terms of the Creative Commons Attribution 4.0 International license (CC-BY 4.0).

https://doi.org/10.5523/bris.3nmqsrbv5ruom2abn0ql6e8yh2

## Acknowledgements

We are grateful to all respondents who completed the questionnaire and to the public contributors for their input into study design and questionnaire development.

## Data and code availability Underlying data

Repository data.bris: The Avian Contact Study: questionnaire data 15 May – 31 July 2024 Due to the sensitivity of the data involved, these data are published as a restricted dataset at the University of Bristol Research Data Repository <u>data.bris</u>, at https://doi.org/10.5523/bris.3nmqsrbv5ruom2abn0ql6e8yh2^35^. The metadata record published openly by the repository at this location clearly states how data can be accessed by *bona fide* researchers. Requests for access will be considered by the University of Bristol Research Data Service, who will assess the motives of potential data re-users before deciding to grant access to the data. No authentic request for access will be refused and re-users will not be charged for any part of this process.

This project contains the following underlying data:

- Data file 1. (Raw underlying questionnaire data – csv file)
- Data file 2. (Raw underlying questionnaire data – .RDS file)
- Data file 2. (Associated data dictionary – csv file)
- Data file 3. (Code for importing underlying data in csv format into R for setting up labelled data – .r file)
- Data file 4. (Blank consent form and participant information sheet – pdf file)

Data are available under the terms of National Archives’ <u>Non-Commercial Government Licence for public sector information</u>.

## Extended data

Repository Zenodo: The Avian Contact Study Questionnaire and Data Dictionary [10.5281/zenodo.13617061]

This project contains the following extended data:

- AvianInfluenzaSocialContactSu.pdf (The final questionnaire REDCap – PDF)
- AvianInfluenzaSocialContactSur_DataDictionary_080824v1.csv (Associated data dictionary –csv file)

Data are available under the terms of the <u>Creative Commons Attribution 4.0 International license</u> (CC-BY 4.0).

### Software availability

Source code available from: https://github.com/amythomas/aviancontactstudy License: <u>Creative Commons Attribution 4.0 International license</u> (CC-BY 4.0).

## Author contributions

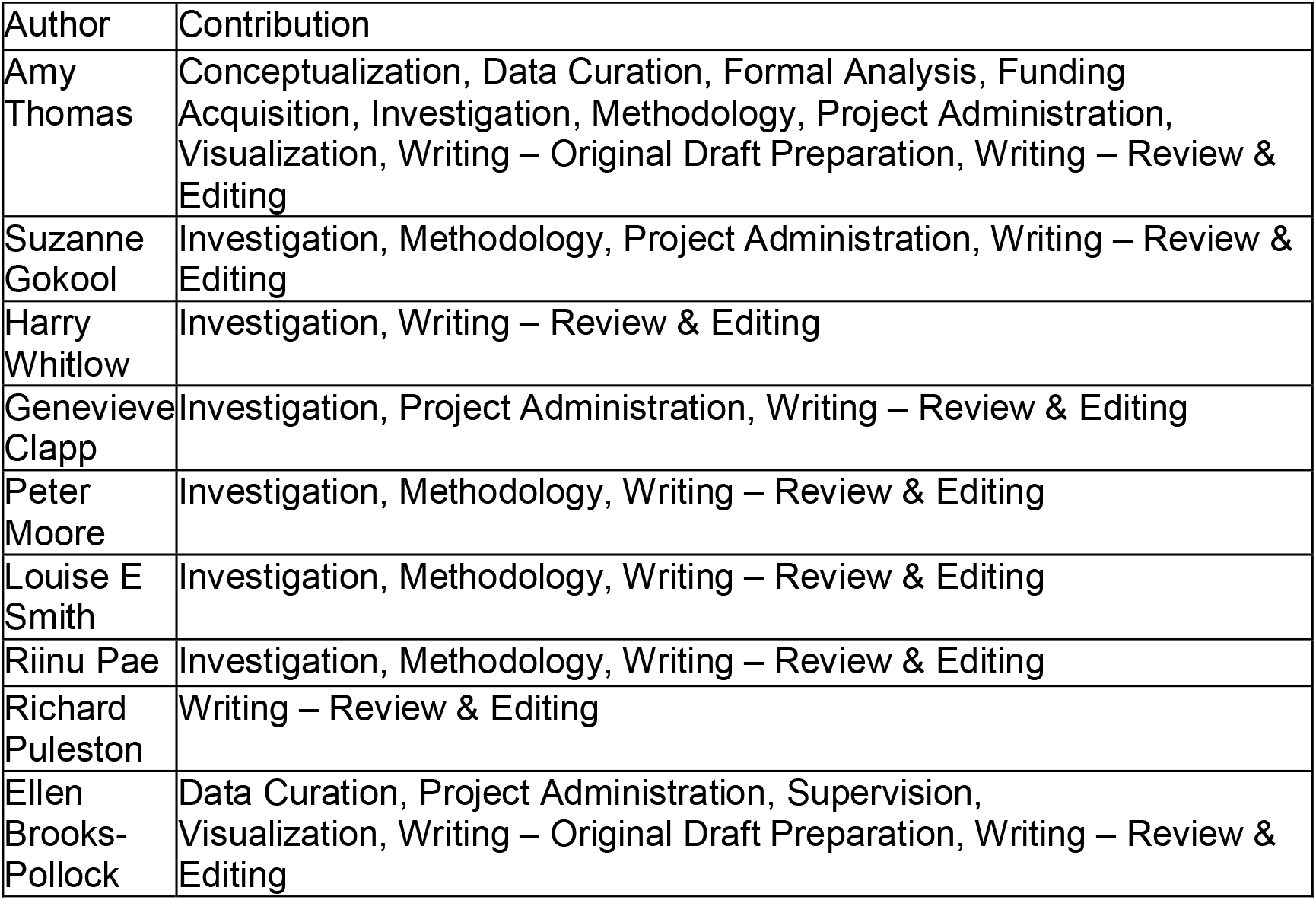

